# Culture-enriched metagenomic sequencing reveals within-patient diversity and transmission of vancomycin-resistant *Enterococcus faecium*

**DOI:** 10.64898/2026.03.11.26348025

**Authors:** Emma G. Mills, Kirsten M. Evans, Ava J. Dorazio, Kevin M. Squires, Alexander J. Sundermann, Madison E. Stellfox, Matthew J. Culyba, Ryan K. Shields, Daria Van Tyne

## Abstract

Colonization of the gastrointestinal (GI) tract by vancomycin-resistant *Enterococcus faecium* (VREfm) often precedes bloodstream infection and serves as a reservoir for onward patient transmission in healthcare settings. Routine clonal isolate-based sequencing often underestimates within-patient diversity, and can miss transmission involving low-abundance and co-colonizing strains. Here we applied culture-enriched metagenomic sequencing to matched GI tract and blood VREfm populations collected ≤14 days apart from 35 patients with positive VREfm blood cultures collected between 2020 and 2025 at a single hospital. GI populations exhibited greater within-patient diversity than bloodstream populations, including multi-strain colonization in five patients. Among single-strain populations, variant analysis suggested distinct environment-specific pressures between the GI tract and bloodstream environments. To assess transmission using culture-enriched metagenomic sequencing, we compared all 70 VREfm populations against 470 contemporary clinical VREfm isolate genomes collected from the same hospital and identified 19 putative transmission clusters, including six clusters involving multi-strain populations. Together, these results demonstrate how culture-enriched metagenomic sequencing improves resolution for assessing within-patient VREfm diversity and enhances the detection of transmission events that could be missed by clonal isolate-based surveillance.

**Impact Statement:** VREfm bloodstream infection is often seeded from bacteria colonizing the gut. The genetic diversity within gut and blood VREfm populations, and the role of this diversity in bacterial transmission, has been difficult to resolve as genomic surveillance typically relies on sequencing a clonal clinical isolate from each patient. Using culture-enriched metagenomic sequencing of matched GI tract and bloodstream VREfm populations from 35 patients at a single hospital, we found that the GI tract reservoir contained VREfm populations with greater strain and variant diversity than populations collected from the bloodstream. By integrating population sequencing with a large collection of VREfm clinical isolate genomes, we further demonstrate that different strains co-colonizing the GI tract of the same patient can reside in multiple putative transmission clusters, revealing potential transmission links that clone-based approaches are likely to miss. These findings demonstrate the potential utility of culture-enriched metagenomic sequencing for higher-resolution hospital surveillance of bacterial transmission. Applying this approach to other bacterial pathogens could improve our ability to detect and interpret transmission involving heterogeneous microbial populations that colonize and infect hospitalized patients.

**Data Summary:** Patient demographic data and clinical characteristics can be found in **Table S1** (online Supplementary Material). All sequencing data generated in this study has been deposited in the National Center for Biotechnology Information (NCBI) under BioProject PRJNA901969, with sample accession numbers listed in **Table S2**. Sequences used to construct the local reference strain database are available at NCBI BioProject PRJNA475751, with accession numbers listed in **Table S3**. Variants identified in single-strain blood and GI populations are listed in **Table S4**. Accession numbers for clinical isolate genomes included in transmission analyses are listed in **Table S5**.

**Repositories:** Sequencing data generated in this study is deposited in the National Center for Biotechnology Information (NCBI) under BioProject PRJNA901969.

## INTRODUCTION

*Enterococcus faecium* is a member of the commensal gut microbiome and is typically detected at low abundance in healthy individuals (1,2). However, healthcare-associated vancomycin-resistant *E. faecium* (VREfm) can cause difficult-to-treat bloodstream infections in immunocompromised and other vulnerable hospitalized patients (1,3). Disruption of the gut microbiota by antibiotic exposure can permit VREfm expansion within the GI tract, thereby increasing the risk of translocation across the intestinal barrier and progression to bloodstream infection (BSI) (1,4–8).

The GI tract is believed to serve as the primary reservoir for invasive VREfm infections, and GI colonization typically precedes bacteremia (4,5,7,8). During hospitalization, patients may acquire and harbor multiple genetically distinct VREfm strains in their GI tract, and the composition of the GI tract VREfm population can shift over time (4,8–13). Consequently, BSIs, including recurrent episodes, may be caused by different strains originating from the same GI reservoir (7,14,15). Beyond serving as a source of infection, GI tract colonization contributes to environmental shedding of VREfm, facilitating transmission within healthcare settings (16–20). Prior studies investigating the diversity and transmission dynamics of GI tract and bloodstream VREfm populations have largely used clone-level sequencing of cultured isolates, which often captures only a single representative colony and might underestimate both the strain-level complexity present and transmission events within these environments (7–10,12,14). As a result, the composition and relative abundance of VREfm strains during colonization and infection are largely unknown, prompting the need for novel approaches capable of resolving strains from diverse populations.

Culture-enriched metagenomic sequencing involves the enrichment of target organisms through selective culturing, followed by high-depth shotgun sequencing of the resulting population (21,22). This approach offers key advantages over traditional clone-based methods by enabling detection of multiple and low-frequency strains, estimation of relative strain abundance, and identification of genetic variation across microbial populations (22–24). This added resolution is particularly important in the context of invasive infections by commensal organisms, where selective pressures likely differ between colonization and infection sites, resulting in environment-specific adaptations (7,25). Culture-enriched metagenomic strategies can therefore provide a more complete picture of genomic variants that may facilitate adaptation to different environmental niches in the host. However, culture-enriched metagenomic approaches also present challenges, including biases introduced during selective culture, uneven strain recovery, and difficulty resolving closely related strains.

In this study, we applied culture-enriched metagenomic sequencing to characterize the diversity of VREfm populations collected from matched GI tract and bloodstream samples from 35 patients at a single hospital. We assessed population diversity at multiple levels (species, sequence type, and strain), quantified the relative abundance of coexisting strains, and identified genetic variation consistent with environment-specific selective pressures. To identify the strains present in GI tract and bloodstream VREfm populations, we constructed a non-redundant representative strain reference database designed to enable sensitive detection and differentiation of closely related VREfm strains within diverse populations. In addition, we demonstrate how culture-enriched metagenomic sequencing can be integrated with existing clinical isolate genomic surveillance methods through the identification of putative multi-strain transmission events. Together these data show that culture-enriched metagenomic sequencing is a useful approach to resolve within-patient diversity and identify putative transmission events within heterogeneous populations of a concerning bacterial pathogen.

## METHODS

### Study Design

This was a retrospective observational study conducted at the University of Pittsburgh Medical Center (UPMC) between June 2020 and January 2025. UPMC is an adult tertiary-care hospital with approximately 750 beds, including 134 critical care beds, and performs more than 400 solid organ transplants annually. Patients were included in the study if they had a vancomycin-resistant *Enterococcus faecium* (VREfm)-positive blood culture and grew vancomycin-resistant enterococci (VRE) from a gastrointestinal (GI) tract sample (either rectal swab or stool), collected within 14 days of the blood culture date. Based on these criteria, 35 patients were included in this study. Patient demographics and clinical characteristics were tabulated by infectious disease physicians by consulting the electronic medical record of each patient (**Table S1**). This study was approved by the Institutional Review Board of the University of Pittsburgh under STUDY20020046.

### Sample processing and sequencing

A single colony from each VREfm-positive blood culture was saved and used as the patient-specific reference isolate, consistent with standard clinical microbiology procedures for bacterial species identification and antimicrobial susceptibility testing. Positive blood cultures were serially diluted and plated onto brain heart infusion (BHI) agar supplemented with 10 µg/mL vancomycin to obtain population plates. Rectal swabs collected from 31 patients were screened for VRE using bile esculin azide (BEA) broth supplemented with 10 µg/mL vancomycin. After overnight growth at 37°C, VRE-positive swabs were identified by black coloration. Positive samples were then serially plated onto BEA agar with vancomycin (50 µg/mL) to generate population plates. Stool samples collected from four patients were suspended in phosphate buffered saline (PBS) and serially plated onto CHROMagar^TM^ VRE and were grown overnight at 37°C. For both GI tract and blood population plates, colonies were pooled to capture approximately 100–1,000 colonies from each population. Genomic DNA was extracted from reference isolates and pooled populations using a DNeasy Blood and Tissue Kit (Qiagen). Patient blood reference isolate and population libraries were prepared using a Nextera library preparation kit and were sequenced on the Illumina NextSeq 2000 platform. Reference isolate genomes were also sequenced on the Oxford Nanopore MinION platform with R9.4.1 flow cells. Libraries were constructed using a rapid multiplex barcoding kit (catalog number SQK-RBK004), and basecalling was performed with Dorado v0.4.1 (Oxford Nanopore Technologies) using default parameters (26). Patient reference genomes were hybrid assembled using Unicycler v0.5.1 (27). Patient sample collection and sequencing accession numbers are accessible in **Table S2**.

### Build of local VRE reference strain database

A non-redundant reference database to assess strain diversity was constructed using all publicly available vancomycin-resistant *Enterococcus* (VRE) clinical isolate genomes, including VREfm and vancomycin-resistant *Enterococcus faecalis* (VREfs) (n = 1,279; 96% VREfm, 4% VREfs), from the Enhanced Detection System for Healthcare-Associated Transmission (EDS-HAT) NCBI BioProject (PRJNA475751) on July 31, 2024 (**Table S3**). Genomes were processed with MOB-suite (mob_recon v3.1.9) (28) to identify and retain contigs >500bp predicted to be chromosomal, thereby excluding plasmid and mobile genetic element-associated sequences that could confound SNP-based distance estimates and strain assignment. Pairwise comparisons of filtered genomes were performed using split kmer analysis (SKA) v1.0 (29), followed by hierarchical clustering using a 100-SNP threshold with average linkage (30). For each cluster, the centroid genome, defined as the isolate with the lowest average pairwise distance to all other genomes in the cluster, was selected as the representative reference genome. If multiple genomes shared identical centroid values, the earliest collected genome was chosen. The clustering and centroid selection process was performed iteratively: after each round, clustered non-centroid genomes were removed, and the remaining centroid and singleton genomes were re-clustered. This process was repeated until all clusters consisted of a single reference genome. The resulting non-redundant representative strain database contained 190 VRE reference genomes, including 160 VREfm and 30 VREfs genomes (**Table S3**).

### Assessment of sample diversity

Species-level composition and potential contamination of isolate and population samples were assessed using Kraken2 v2.1.3 (31) (**Table S2**). Reads assigned to non-enterococcal species were removed with KrakenTools (32). Population sequencing reads were then compared against the local VRE reference strain database described above using TRACS v1.0.1 to determine sequence type and reference strain assignment (33). Sequence types were assigned based on the sequence type of the TRACS reference strain identified. Strain identification was performed using TRACS, with strain presence defined as an F-match score of ≥0.9 and unique intersect base pairs ≥100,000. These thresholds were selected to detect low-abundance strains while maintaining specificity for true strain matches. All identified strains had a ‘FALSE’ designation for the Potential False Negative flag, indicating sufficient sketch sizes and reliable average nucleotide identity estimation.

Approximate strain abundances for multi-strain populations were estimated by mapping population reads to their corresponding strain references using QIAGEN CLC Genomics Workbench v24.0 (https://digitalinsights.qiagen.com/). Population assemblies were generated using SPAdes v4.0.0 (34,35) with the --meta flag, and assemblies were annotated with Prokka v1.14.5 (36) to examine the relationship between sample diversity and genome size (**Table S2**).

### Variant identification

Variant analysis was performed on all 35 blood populations as well as the 26 single-strain GI tract populations that matched the strain identified for the blood reference isolate from the same patient. For these 61 samples, population sequencing reads were mapped to the corresponding patient-specific reference genome using breseq v0.39.0 (37) in population mode with default settings. Variants were retained if they met the following criteria: i) located on the chromosome, ii) supported by ≥20X coverage, and iii) present at ≥10% allele frequency. To minimize the influence of recombination, regions containing more than two variants within a 1,000 bp window were excluded from downstream analysis. Identified variants are listed in **Table S4**.

### Functional and gene enrichment of mutated genes

Genes with nonsynonymous single nucleotide polymorphisms (SNPs) or insertions/deletion variants within the coding region were assigned Clusters of Orthologous Genes (COG) functional categories based on Prokka v1.14.5 (36) and eggNOG-mapper v5.0 (38,39) annotations (**Table S4**). Genome-wide COG assignments were generated from the chromosome of a representative ST117 isolate (1558) to serve as a reference for COG category distribution. For genes annotated with multiple COG categories, all assigned categories were included in the analysis. Functional enrichment was assessed by comparing the distribution of COG categories among all mutated genes, genes mutated among GI tract samples, or among bloodstream samples, to the genome-wide COG distribution of the representative ST117 isolate 1558 (25). Gene-level enrichment was evaluated by comparing the observed versus expected numbers of genes mutated once, twice, or three or more times (40). The expected distribution was generated by simulating 10,000 permutations of the observed mutations, deduplicated by patient (n = 119 mutations), randomly assigned across the pangenome identified by Roary (41) (n = 4670 chromosomal genes), factoring in gene length and total mutations. Empirical p-values were calculated by comparing observed mutation counts against the simulated distribution. Amino acid sequences from hypothetical genes that were mutated two or more times were further investigated using BLASTp to identify conserved protein domains (42,43).

### Comparative genomics of population samples and clinical isolates

TRACS v1.0.1 (33) was used to compare population samples to VREfm clinical isolates collected by EDS-HAT (BioProject PRJNA475751) during the study period plus an additional 3 months before and after the study sampling window (March 2020 – April 2025) (**Table S5**). Clinical isolates were first deduplicated by patient and sequence type and were then evaluated for inclusion against the local reference strain database. Clinical isolates were included in downstream comparisons if a corresponding database reference met the following inclusion criteria: (i) matching sequence type between the reference strain and the clinical isolate, (ii) the F-match score was ≥0.9, and (iii) the number of unique intersect base pairs was ≥100,000. Clinical isolate sequence types were determined using mlst v 2.11 with the PubMLST database (44,45). This resulted in 470 unique clinical isolates with high confidence matches to strains in the reference database.

TRACS was then used to directly compare all 470 clinical isolates against blood and GI tract population samples from the 35 study patients. Recombination was filtered using the sliding-window approach implemented in TRACS (--filter flag), and putative transmission clusters were identified using hierarchical clustering with a threshold of ≤10 SNPs and average linkage (30,33). Clusters were visualized using Gephi v0.10.1 (46).

### Statistical Analyses

A chi-square test for independence (α = 0.05) was used to evaluate whether the proportion of GI tract samples collected after the positive blood culture date was associated with sample diversity, defined as single-strain, different-strain, or multi-strain populations. To quantify differences in variant counts between paired GI tract and blood populations from the 26 patients with matching single-strain samples, a paired two-sided Wilcoxon signed-rank test was performed (α = 0.05). Linear regression was used to evaluate the relationship between sampling interval and the absolute number of variant differences between each patient’s GI tract and blood populations. Enrichment of functional COG categories was determined using Fisher’s Exact Test (25).

## RESULTS

### Study design and patient demographics

Between June 2020 and January 2025, we identified 35 patients at UPMC with VREfm-positive blood cultures who also had a VRE-positive GI tract sample collected up to 14 days before or after their positive blood culture. Patients tended to be male (71%, n = 25) and were generally older, with a median age of 62 years (range 32 - 87) (**Table 1, Table S1**). Prior solid organ transplantation (34%), malignancy (31%), and immunosuppression (29%), all known risk factors for VREfm bloodstream infection (BSI), were common among patients in this cohort (7,47). GI translocation was the most frequently suspected source of BSIs (54%), followed by intra-abdominal/hepatobiliary infection (37%), and central line-associated BSI (CLABSI) (29%). The duration of positive blood cultures was generally short, with most resolving within one day; however, BSI persisted longer than seven days in four patients, including one case that lasted 19 days.

**Table 1.** Clinical characteristics of patients with vancomycin-resistant *E. faecium*-positive blood cultures.

| Characteristic | Overall (n = 35) |
| --- | --- |
| **Age, years: median (range)** | 62 (32 – 87) |
| **Sex, n (%)** |  |
| Male | 25 (71%) |
| Female | 10 (29%) |
| **Comorbidities, n (%)** |  |
| Solid Organ Transplant | 12 (34%) |
| Malignancy | 11 (31%) |
| Immunosuppression | 10 (29%) |
| Diabetes mellitus | 9 (26%) |
| **Suspected Source, n (%)** |  |
| GI translocation | 19 (54%) |
| Intra-abdominal / hepatobiliary infection | 13 (37%) |
| CLABSI | 10 (29%) |
| **Duration of Positive Blood Culture, days: median (range)** | 1 (1 – 19) |

\*Probable sources of positive blood cultures were assigned based on clinical and microbiologic evaluation. Some patients had multiple probable sources of positive blood cultures and multiple comorbidities. GI = gastrointestinal, CLABSI = central line-associated bloodstream infection.

### VRE GI tract populations harbor greater genetic diversity than bloodstream populations

We performed culture-enriched metagenomic sequencing of matched GI tract and bloodstream populations collected from each patient to capture population-level vancomycin-resistant *Enterococcus* (VRE) diversity. Colonies that grew on VRE-selective agar were pooled (100-1000 colonies from each plate) and sequenced at high coverage (median 546x). We investigated population diversity at three levels: species, multi-locus sequence type (ST), and strain. Starting with species-level diversity, we used Kraken2 to approximate species abundance (31). Consistent with the identification of VREfm in all 35 blood cultures, all blood populations contained only VREfm. Nearly all GI tract populations also contained only VREfm, with one patient’s GI population showing the presence of both VREfm (64%) and vancomycin-resistant *E. faecalis* (VREfs) (36%) (**Table S2**). These findings are consistent with the higher VREfm burden at our center compared to VREfs (19,20).

To next investigate both ST and strain level diversity, we used the bioinformatics tool TRACS (33), which relies on a reference genome database to identify and compare strains present in metagenomic samples. To capture the local diversity of VREfm and VREfs strains at our center, we constructed a non-redundant reference strain database using genomes collected between 2016 and 2019 through the Enhanced Detection System for Healthcare-Associated Transmission (EDS-HAT) surveillance program (**Table S3**) (48). Briefly, genomes were compared using split kmer analysis (SKA) (29) and were iteratively deduplicated using a 100-SNP cut-off. The final database included 190 genomes (160 VREfm, 30 VREfs) spanning 61 STs (**Table S3**). Across all 35 patients, no ST-level diversity was detected among blood VREfm populations (**Fig 1, Table S2**). Most patients (n = 26, 74%) had matching, single-ST populations in their blood and GI tract samples, most commonly ST117 (n = 15, 43%) (**Fig. 1A**). Four patients (11%) carried different STs in their blood and GI tract populations, suggesting that the BSI may not have arisen from the GI reservoir, or that the colonizing strain was missed due to timing or method of sampling (**Fig. 1B**). Most interestingly, five patients (14%) had multiple STs detected in their GI samples, including one patient with three different STs (VREfm ST412, VREfm ST117, and VREfs ST6) (**Fig. 1C**). These multi-ST populations demonstrated higher gene content and larger genome sizes compared to single-ST populations, consistent with co-colonization by multiple genetically distinct VRE lineages (**Fig S1, Table S2**). To approximate ST abundance in the multi-ST populations, we stringently mapped metagenomic reads to the corresponding TRACS-identified reference genomes. In four out of the five patients with multi-ST populations, the ST at the highest frequency in the GI tract was also the ST present in the blood population (**Fig. 1C, Table S2**). In the fifth patient, the GI tract population comprised a near-equal mix of ST17 (43.4%) and ST1478 (56.6%) strains, while only ST17 was recovered from the bloodstream. For the 31 patients with matching sequence types across GI tract and bloodstream populations, the same strain was consistently identified in both the GI tract and the bloodstream, indicating that bloodstream infections typically arise from the dominant GI tract strain (**Fig. 1A and 1C, Table S2**) (4,50).

**Figure 1.**
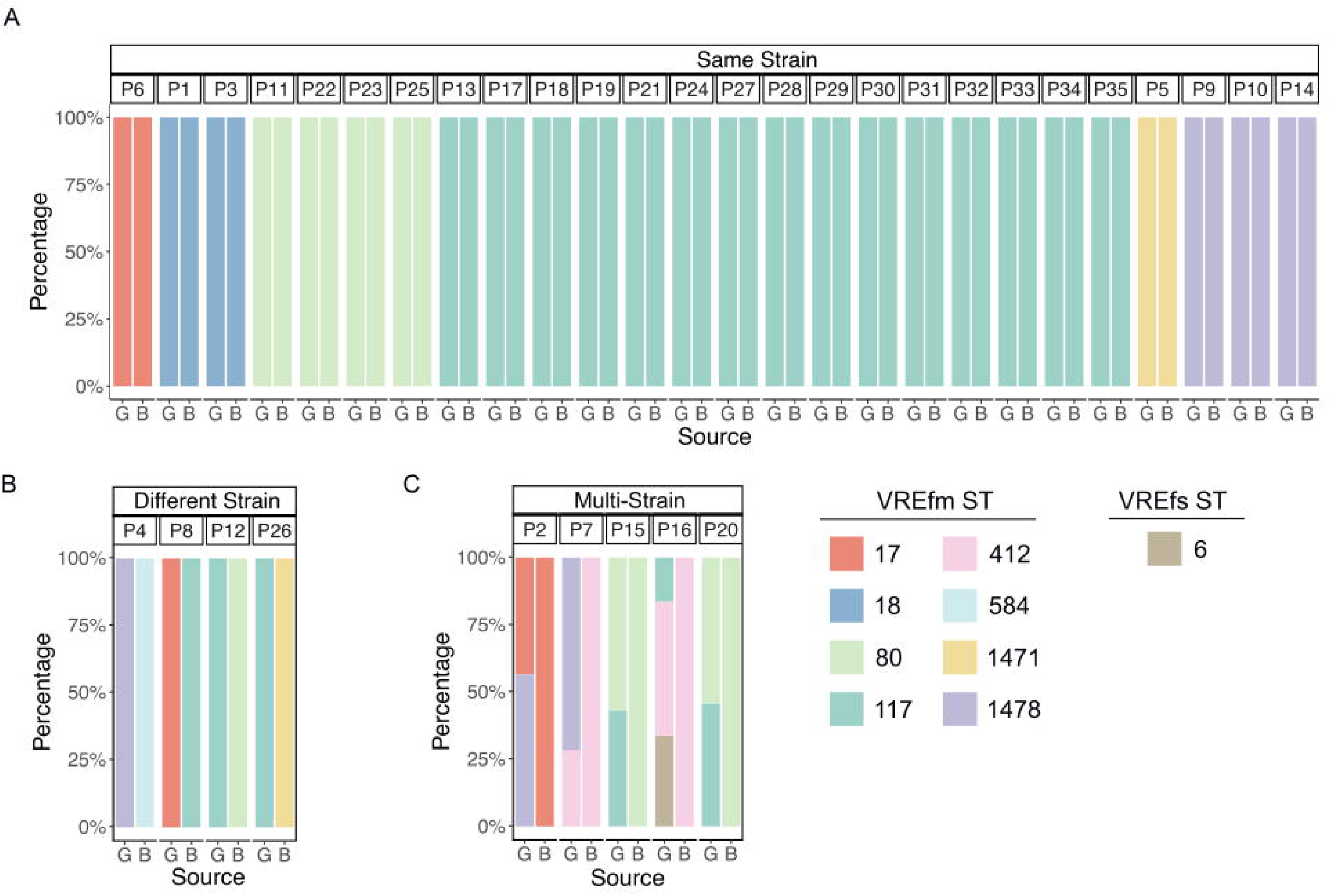
Within-patient diversity of matched gastrointestinal tract and bloodstream samples collected from 35 patients with VREfm BSI. Each patient’s gastrointestinal tract (G) and blood (B) populations were plated onto, pooled, and sequenced from vancomycin-resistant *Enterococcus* selective agar. A) Within-patient populations consisting of the same, singular strain. B) Within-patient populations consisting of different, singular strains. C) Within-patient populations consisting of multiple strains. The bioinformatics tool TRACS identified the strains present in each sample using a custom local and non-redundant reference database. Gastrointestinal tract multi-strain populations were stringently mapped against the reference identified by TRACS using CLC Genomic Workbench to approximate relative strain abundance. Panels follow the same color-coding scheme for vancomycin-resistant *Enterococcus faecium* (VREfm) and *Enterococcous faecalis* (VREfs) sequence types (ST) as shown.

We assessed whether timing or order of sampling were associated with different strain and multi-strain patients (**Fig. 2A**). We found no evidence of such an association (p = 0.805, χ² test for independence), suggesting that the timing of GI sampling did not meaningfully influence the observed strain diversity in this cohort. To further resolve within-patient diversity, we next investigated variant-level differences in the 26 patients with matching single-strain GI tract and bloodstream populations by identifying mutations at greater than 10% frequency in each population compared with the blood isolate reference genome from each patient (**Fig. 2B**). GI tract populations had significantly more variants than the corresponding blood populations (p = 0.0003). We then assessed whether the number of variants was associated with the time that passed between GI tract and blood sampling. We calculated the absolute variant differences for each patient and found no association with sampling interval (**Fig. 2C**, p = 0.92, linear regression), suggesting that the greater diversity we observed in the GI tract is likely due to biological factors such as a larger effective population size, and/or variable ecological niches.

**Figure 2.**
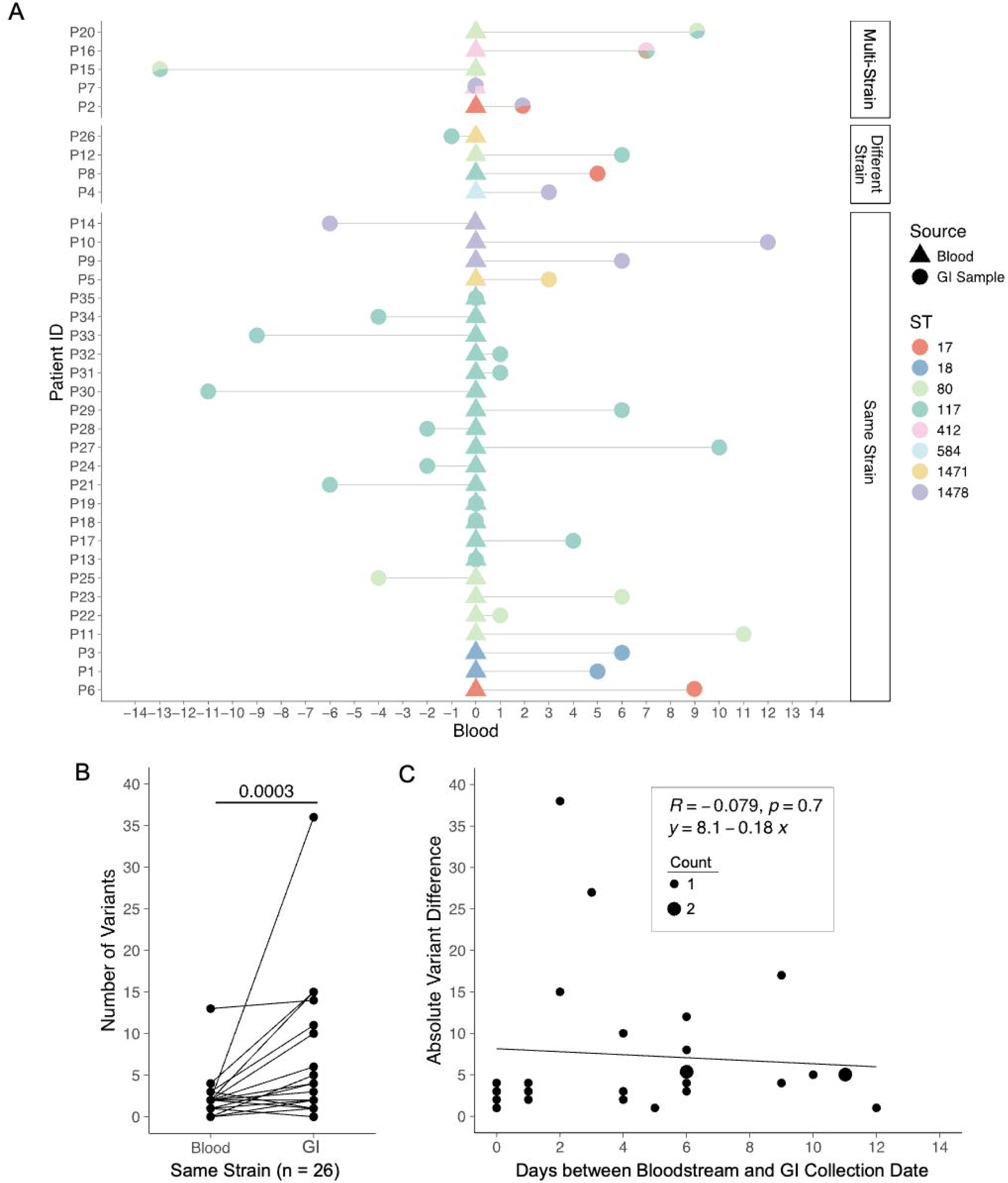
Gastrointestinal VREfm populations show greater genetic diversity than bloodstream populations, independent of sampling interval. A) Population diversity by sampling timeframe. Each patient’s GI tract sample collection date is shown in relation to the blood culture date, centered at zero. Patients are grouped by whether their blood (triangle) and GI tract (circle) populations have the same, different, or multiple strains. B) Variant diversity between blood and GI tract populations. For the 26 patients with matching, single-strain populations, GI tract and bloodstream populations were compared to patient-specific hybrid assemblies using *breseq;* chromosomal variants were included in the analysis. A paired two-sided Wilcoxon signed-rank test compared the number of variants detected in bloodstream versus GI tract populations (p = 0.0003). C) Linear regression comparing the absolute variant differences between GI tract and bloodstream populations vs. days between sampling for 26 patients with matching single-strain populations. Count indicates the number of datapoints at each position.

Overall, these findings demonstrate that at the species, strain, and variant levels, GI tract populations of VREfm show greater diversity than bloodstream populations.

### Functional enrichment and repeatedly mutated genes across single-strain populations

To investigate whether the bloodstream and GI tract environments select for different genetic functions, we evaluated whether variants present in populations sampled from these two sites were enriched in particular functional pathways, operons, or genes. We identified variants present at greater than 10% frequency in all single-strain bloodstream populations (n = 35) and the 26 matched single-strain GI tract populations, totaling 61 populations. Variants were identified by mapping sequences to patient specific-reference genomes, and only variants identified on the chromosome and encoding nonsynonymous or insertion/deletion mutations within coding regions were considered (**Table S4**).

To determine whether different categories of genes were mutated among GI tract and bloodstream populations, we first assigned Clusters of Orthologous Genes (COG) categories to all genes with nonsynonymous or insertion/deletion variants among GI tract and bloodstream populations. We then compared the distribution of COG categories among GI tract, bloodstream, and shared site variants against the distribution of COG categories in a representative whole genome using Fisher’s Exact Test (**Fig. 3A, Table S4**). We identified enrichment of COG category M, including genes involved in cell wall/membrane/envelope biogenesis, among the variants detected in populations from both sites; however, enrichment was stronger among GI tract populations (GI+Blood: p = 0.0153; GI only: p = 0.0032). Genes in this category included peptidoglycan synthesis and cell envelope-modifying genes (*murC*, *murG*, *pbpF*, *wecA*, *mprF*) (**Table S4**), which could indicate an accumulation of mutations in response to cell surface stress. Similarly, enrichment of COG category X, including genes in the mobilome, was identified among the variants detected in populations from both sites, but again enrichment was stronger among GI tract populations (GI+Blood: p = 0.0076; GI only: p = 0.0096), perhaps reflecting increased mobile genetic element activity in this environment.

**Figure 3.**
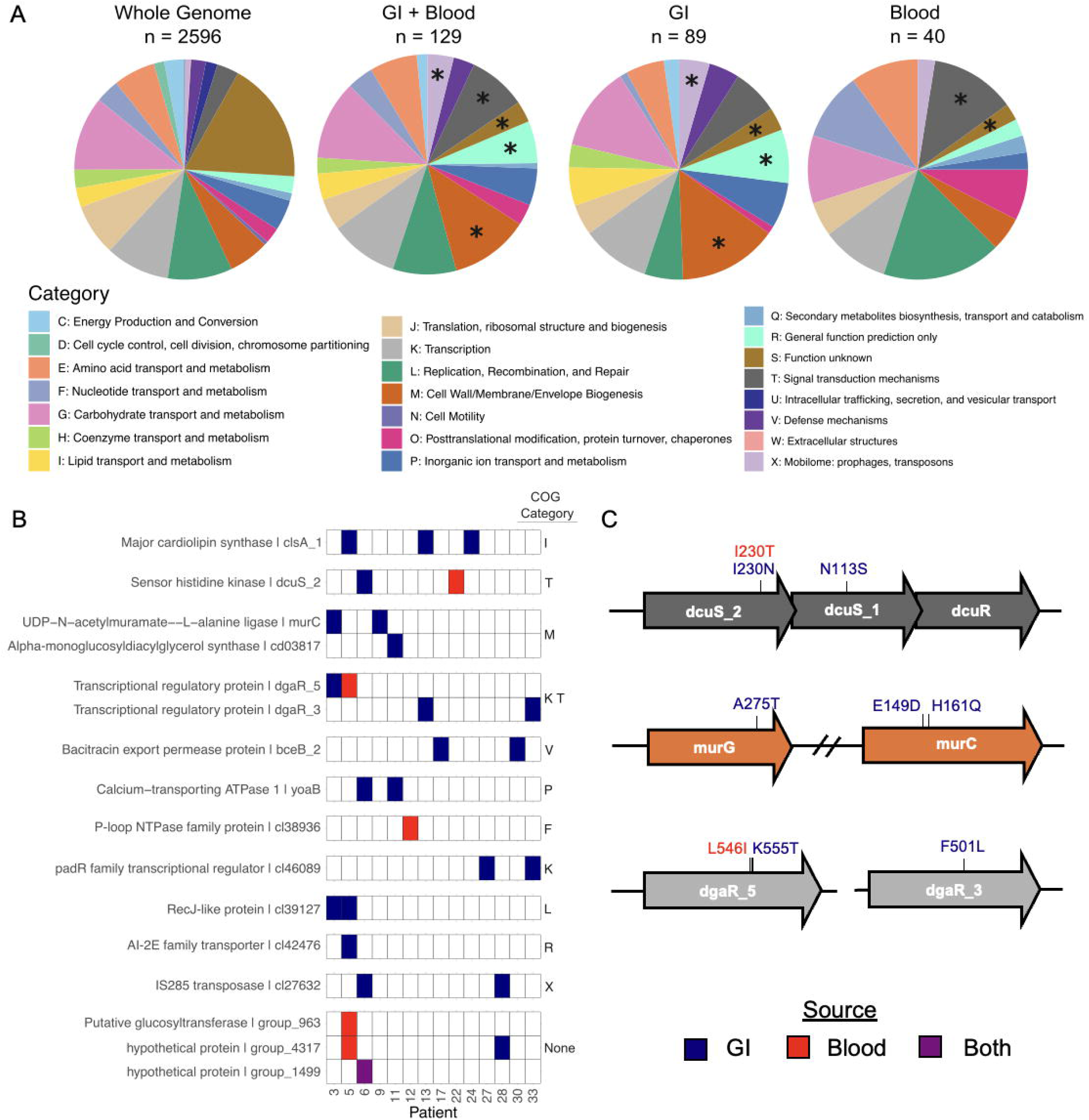
Enrichment of functional categories and recurrently mutated genes identified in single-strain populations. Patient GI tract and blood populations consisting of a singular strain that matched the respective patient-specific bloodstream reference (n = 61) were compared using breseq. Only nonsynonymous, nonsense, and indel variants within chromosomal coding regions were included. A) Enrichment of COG-predicted functional categories among all mutated genes. Enrichment was assessed by comparing the distribution of COG categories identified among mutated genes overall, in GI tract populations, or in bloodstream populations, to the genome-wide distribution using Fisher’s exact test (p ≤ 0.05*). B) Genes mutated two or three times, grouped by COG functional category and patient. Hypothetical proteins are noted by conserved protein domain and Roary gene group. C) Representative operons and gene clusters containing multiple independent mutations or genes with related functions. Genes are colored by associated COG functional category. In panels B and C, blue indicates mutations identified in GI tract populations; red indicates mutations identified in blood populations, and purple indicates mutations identified in both.

Finally, COG category T, including genes involved in signal transduction, was enriched among variants identified in populations from both sites but enrichment was stronger among blood populations (GI+Blood: p = 0.0045; Blood only: p = 0.0101). Genes in this category included two-component systems and associated transcription regulators (*dcuS*, *dagR*, *manR*). Together, these patterns suggest differential selective pressures in the GI tract and bloodstream environments.

Next, we examined whether specific genes or loci might be under selection, as evidenced by the presence of repeated and independent mutations. We identified 16 genes with repeated mutations, including *clsA,* a daptomycin resistance–associated gene, that was mutated in GI tract populations in three patients (**Fig. 3B**) (51). Additionally, the number of genes mutated twice was significantly greater than expected by chance (p < 0.0001). When considering repeated mutations at the locus or operon level, we observed multiple mutations in the *dcuS*–*dcuR* two-component sensing system locus, which regulates C4-dicarboxylate metabolism (52). Two populations had unique amino acid variants at the same residue (I230T/N) in *dcuS_2*, while a third population carried a distinct mutation in *dcuS_1* (**Fig. 3C**). We also identified mutations in the *mur* operon (involved with peptidoglycan synthesis) in the GI tract populations of two patients, with *murC* mutated in both patients and one of these patients also harboring a mutation in *murG* (**Fig. 3C**) (53). Additionally, we detected mutations in two sigma 54-like transcription regulators, here named *dgaR_5* and *dgaR_3*. In *dgaR_5*, a regulator located upstream of a putative mannose and sorbose phosphotransferase systems (PTS), we identified a mutation in the bloodstream population of one patient and in the GI tract population of another patient. Further, we observed the same mutation (F501L) in *dgaR_3,* a regulator located upstream of a putative lichenan and cellobiose PTS, in the GI tract populations of two different patients (**Fig. 3C**). Together, these findings indicate that while a relatively small number of genes were repeatedly mutated in GI tract and bloodstream populations, some evidence of parallelism was still apparent, suggesting environment-specific adaptations across patients.

### Multi-strain populations form independent clusters with clinical isolates

We next asked whether culture-enriched metagenomic sequencing could be integrated with routine genome sequencing surveillance of clonal clinical isolates to improve detection of putative VREfm transmission. We compared all blood and GI tract populations to 470 unique patient isolates collected during our study period, plus the three months before and after, through EDS-HAT (48). Clinical isolates and populations were clustered using TRACS with a ≤10-SNP threshold (**Fig. 4**). This analysis identified 13 putative transmission clusters consisting of single-strain populations and clinical isolates, involving 2 to 14 patients per cluster. When we examined populations containing multiple strains, we found six transmission clusters in which multi-strain populations clustered with clinical isolates, involving 2 to 15 patients per cluster.

**Figure 4.**
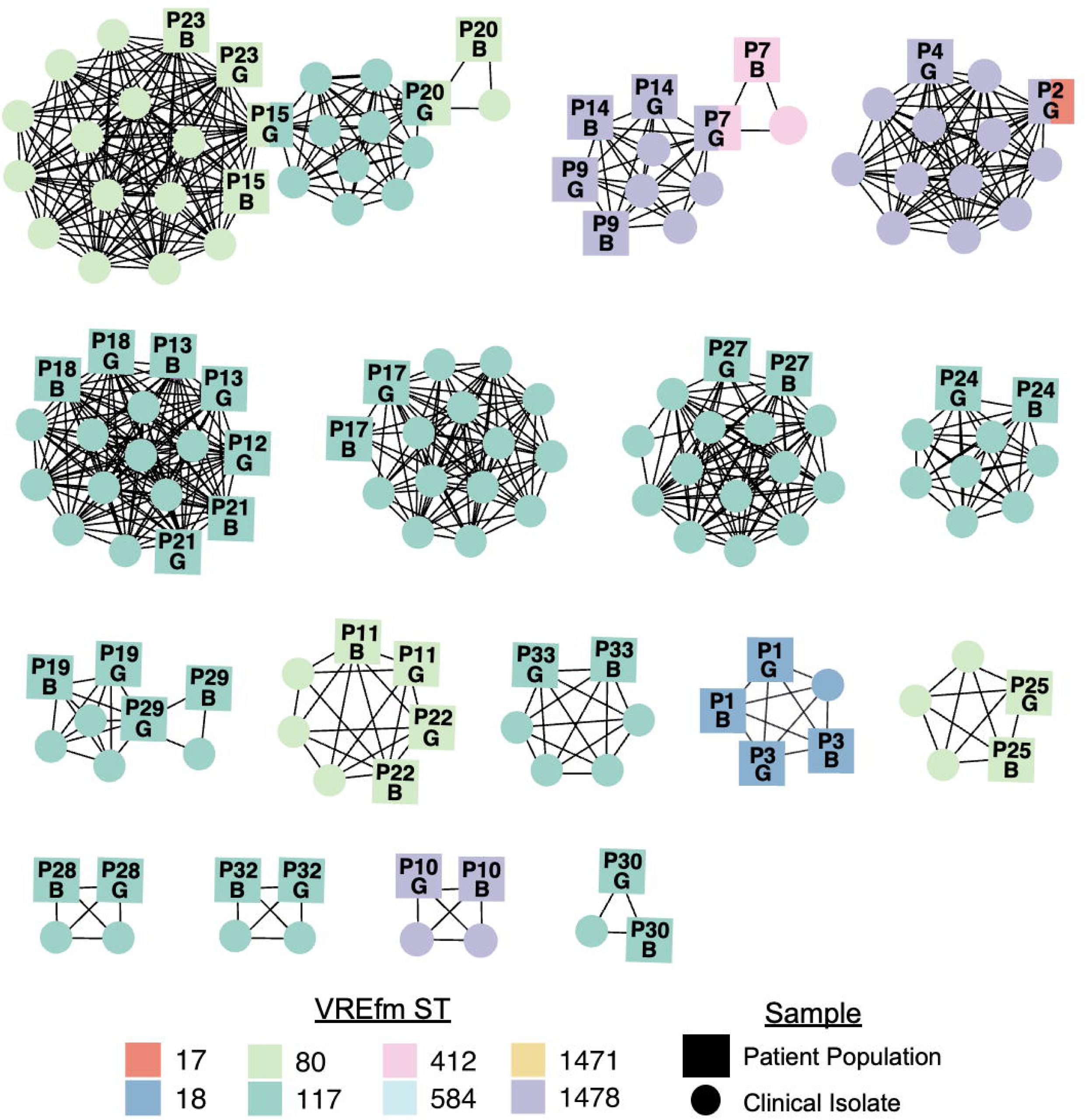
Multi-strain VREfm populations clusters with clinical isolate genomes. Population-level clustering of VREfm populations with clinical isolate genomes. Blood (B) and gastrointestinal tract (G) patient populations (squares) were compared using TRACS with 470 clinical VREfm isolate genomes (circles) collected contemporaneously. Genetically related isolates (≤10 SNPs) are connected by a black line. Samples are colored by sequence type (ST).

Notably, three patients with multi-strain GI tract populations each resided in two separate putative transmission clusters. If we had only sampled a single isolate from each patient, the presence of these patients in two different clusters would not have been apparent. Collectively, these findings demonstrate that culture-enriched metagenomic sequencing complements existing clone-level surveillance and enhances the resolution of VREfm transmission tracking within healthcare settings.

## DISCUSSION

In this study, we used culture-enriched metagenomic sequencing to characterize within-patient diversity and transmission dynamics of VREfm across matched gastrointestinal and bloodstream populations collected from hospitalized patients. This approach revealed substantially greater VREfm diversity in the GI tract compared to the bloodstream, including multi-strain populations that would have been missed by traditional clone-level sequencing.

Variant-level analyses identified recurrently mutated genes and functional pathways, supporting the idea that distinct environment-specific pressures exert selection on VREfm populations during colonization and infection. Finally, by integrating metagenomic data with 470 clinical isolates from the same hospital, we showed that multi-strain populations can reside in multiple independent transmission clusters, demonstrating that culture-enriched metagenomic sequencing provides enhanced resolution for surveillance and can detect transmission events that would be overlooked by standard isolate-based methods.

Understanding the population-level diversity of VREfm within hosts is critical, as variation among coexisting strains can shape infection risk, therapeutic failure, and opportunities for onward transmission in healthcare settings. Gastrointestinal VREfm populations likely exhibit greater genetic diversity than bloodstream populations because the gut supports far larger populations, greater ecological complexity, heterogeneous selective pressures, and continual exposure to new VREfm strains from the hospital environment (4,8,12,54). Studies have shown that VREfm GI tract colonization can sometimes reach very high densities (≥90% of the GI consortia), creating ample opportunity for within-host diversification during prolonged hospitalization and fluctuating antibiotic exposure (4,12). In contrast, bloodstream population sizes are likely much smaller due to the strong bottleneck imposed during translocation from the GI tract into the bloodstream coupled with nutritional, antibiotic, and immune selection. Prior studies have shown that bloodstream isolates often represent one of several coexisting GI tract lineages, and that recurrent infections may arise from previously low-abundance or newly acquired strains in the gut (8,14). Consistent with these findings, we observed markedly greater diversity among GI tract VREfm populations, including multi-strain populations, whereas bloodstream samples uniformly contained a single strain. We also identified substantially more variants among GI tract VREfm populations, further indicating that only a subset of GI variants successfully translocate and establish infection in the bloodstream.

Our analysis also revealed recurrent mutations across several functional pathways and genes, suggesting convergent adaptation to host-specific selective pressures. Mutations affecting cell wall and cell envelope biogenesis were enriched among GI tract VREfm populations, consistent with prior work showing frequent peptidoglycan and cell envelope remodeling during intestinal colonization (8,12). We also observed enrichment of mutations in metabolic sensing and carbohydrate utilization pathways, suggesting that nutrient availability shapes adaptation in both the GI tract and the bloodstream, which has also been reported previously (8,25). Consistent with a prior study by Chilambi *et al*., bloodstream populations showed an increased presence of mutations in nucleotide biosynthesis pathways, although this trend in our data did not reach statistical significance (25). Notably, multiple patients in our study carried mutations in *clsA*, a cardiolipin synthase associated with daptomycin resistance, highlighting the potential role of antibiotic pressure in shaping within-patient evolution (51,55,56). The occurrence of multiple mutations in these functional categories across patients suggests that shared selective forces, nutrient availability, host-derived stressors, and antimicrobial exposure all drive parallel evolutionary trajectories during VREfm colonization and infection.

Our healthcare system performs real-time genomic surveillance of clinical isolates from healthcare-associated infections (20,48), offering a unique opportunity to integrate our results with an ongoing genomic surveillance program at the same hospital. Prior studies have shown that VREfm is a major pathogen involved in widespread transmission at our center (19,20,48,49,57). This high VREfm burden is not unique to our hospital, as numerous healthcare systems worldwide have reported rampant transmission of VREfm within healthcare settings (58–63). In this study, culture-enriched metagenomic sequencing enabled the detection of putative transmission events involving multi-strain GI tract populations, identifying several instances of multi-strain transmission that would likely be missed by single-colony approaches. Given the established role of the hospital environment as a reservoir for VREfm exposure and transmission (64–67), our culture-enriched metagenomic sequencing approach is well suited for environmental surveillance, as it allows pooled analysis of entire microbial populations rather than reliance on a single isolate, increasing throughput and reducing the risk of missing transmitting strains. Importantly, this approach is also applicable to other healthcare-associated pathogens characterized by polyclonal colonization, infection, and environmental persistence (22,68–71).

In performing this study, we evaluated several metagenomic strain tracking tools and ultimately selected TRACS as our preferred method to identify strain presence and putative transmission events after finding that other tools had shortcomings that hindered their use. StrainGE, while effective for studies with fewer samples, was substantially more computationally intensive at the scale of our study, and its average callable identity nucleotide (ACIN) metric was less intuitive to interpret than recombination-filtered SNP distances for assessing relatedness between strains (72). In contrast, TRACS provides SNP-based comparisons and includes a recombination-masking step, which is particularly important for highly recombinogenic *E. faecium* genomes (73). We also evaluated metagenomic binning approaches like MetaBAT 2 and CONCOCT, which reliably distinguished VREfm from VREfs but lacked sufficient resolution to separate closely related strains within each species (74,75). Similarly, StrainPhlAn enabled population-level phylogenetic comparisons but primarily captured the dominant strain within a sample, limiting its ability to detect lower-frequency strains involved in transmission (76). Together, these considerations motivated our use of TRACS with a curated, non-redundant reference database, enabling scalable integration of metagenomic samples with clinical isolates for putative transmission cluster identification.

This study has several limitations. First, GI tract samples were collected using rectal swabs or stool samples, which capture only a small fraction of the microbial community and may underrepresent sample diversity. Similarly, blood culture collection imposes a substantial population bottleneck, as only a small number of circulating bacteria are sampled and subsequently expanded *in vitro*. Second, because samples were collected at a single timepoint relative to the bloodstream infection, our data provide only a snapshot of within-patient diversity, and we cannot determine how these populations fluctuate over time or how dynamics preceding sampling may have shaped our observations. Third, transmission analyses were conducted without full epidemiologic data, limiting our ability to infer directionality or confirm patient-to-patient spread. Fourth, the study was performed at a single center with a relatively small cohort, and all patients were included on the basis of having a positive VREfm blood culture, meaning that we did not include any patients with asymptomatic GI tract colonization. Lastly, the TRACS reference database we used was constructed using draft genome assemblies. While closed genomes represent the best practice for reference-based analyses, generating closed genomes at the scale required to capture strain-level diversity was not feasible. Nonetheless, our findings demonstrate the utility of a culture-enriched metagenomics approach for resolving strain-level diversity and identifying putative transmission events of diverse VREfm populations, which could be applied broadly to other nosocomial pathogens.

In conclusion, here we used culture-enriched metagenomic sequencing to attain a more complete view of within-patient VREfm population diversity by capturing more species, ST, strain and variant level variation than would be possible with clone-based sequencing. This approach enabled detection of multi-strain populations and low-frequency variants, revealing site-specific selective pressures on GI tract and bloodstream populations of VREfm. Further, this approach could be readily integrated with routine clinical isolate surveillance to uncover transmission events involving polyclonal pathogen populations. These findings demonstrate that culture-enriched metagenomics is a scalable and broadly applicable strategy for surveillance of VREfm and potentially other healthcare-associated pathogens across diverse patient populations and environmental reservoirs.

## Data Availability

Patient demographic data and clinical characteristics can be found in Table S1 (online Supplementary Material). All sequencing data generated in this study has been deposited in the National Center for Biotechnology Information (NCBI) under BioProject PRJNA901969, with sample accession numbers listed in Table S2. Sequences used to construct the local reference strain database are available at NCBI BioProject PRJNA475751, with accession numbers listed in Table S3. Variants identified in single-strain blood and GI populations are listed in Table S4. Accession numbers for clinical isolate genomes included in transmission analyses are listed in Table S5.

## Author Contributions

E.G.M. and D.V.T. designed the study. E.G.M., K.M.E., A.J.D., K.M.S., A.J.S., M.E.S., R.K.S., and D.V.T. collected data. E.G.M., M.J.C., and D.V.T. analyzed and interpreted the data. E.G.M. and D.V.T. drafted the paper and figures. All authors reviewed and edited the paper.

## Ethical Approval

Ethical approval for this study was obtained from the University of Pittsburgh Institutional Review Board (STUDY20020046).

## Consent for Publication

Not applicable, as this manuscript does not contain any identifiable individual data.

## Acknowledgements

We would like to thank all members of Microbial Genomics Laboratory at the University of Pittsburgh, and in particular Drs. Lee Harrison and Lora Pless, for their efforts in creating and maintaining the EDS-HAT surveillance system.

---

**Figure S1.**
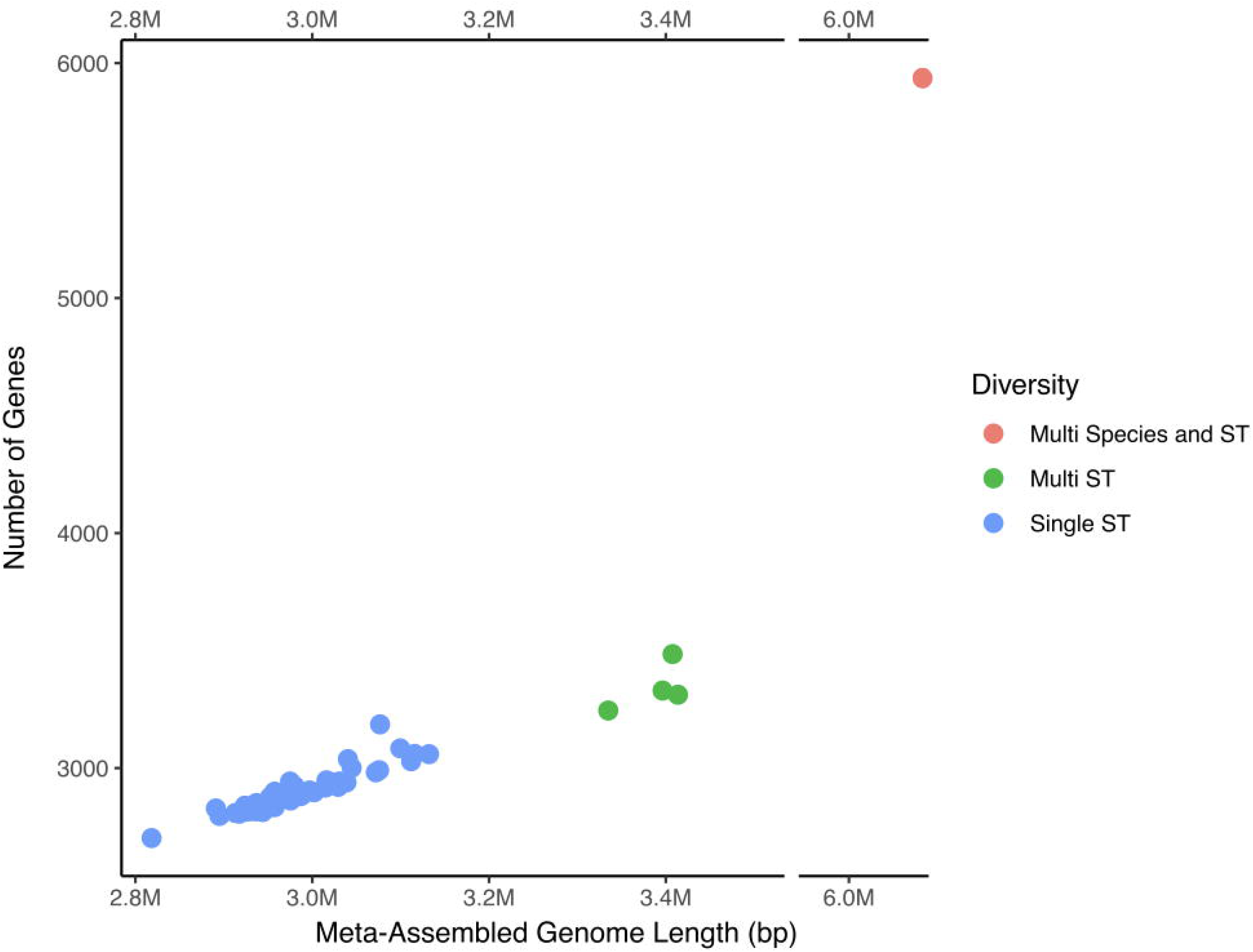
Populations with greater diversity show larger metagenome-assembled genomes and greater gene content. Blood and gastrointestinal tract populations were assembled using SPAdes with the --meta flag to generate metagenome-assembled genomes (MAGs). Gene counts were tabulated using Prokka. Each point represents one population, colored by within-sample diversity: multi-species and multi-sequence type (ST) populations (red), multi-ST populations (green), and single-ST populations (blue).

